# Detection of Lumpy Skin Disease Virus Reads in the Human Upper Respiratory Tract Microbiome Requires Further Investigation

**DOI:** 10.1101/2024.04.22.24306117

**Authors:** Siddharth Singh Tomar, Krishna Khairnar

## Abstract

Lumpy skin disease virus (LSDV), a double-stranded DNA virus from the Capripoxvirus genus, primarily affects Bos indicus, Bos taurus breeds, and water buffalo. Arthropod vectors, including mosquitoes and biting flies, are the main LSDV transmitters. Although LSDV is not zoonotic, this study unexpectedly detected LSDV reads in the upper respiratory tract microbiome of humans from rural and urban areas in Maharashtra, India. Nasopharyngeal and oropharyngeal swab samples collected for SARS-CoV-2 surveillance underwent whole-genome metagenomics sequencing, revealing LSDV reads in 25% of samples. SKA analysis provided insights into sample relatedness despite the low coverage of LSDV reads with the reference genome. Our findings, which include the detection of LSDV contigs aligning to specific locations on the reference genome, suggest a common source for LSDV reads, potentially shared water sources or milk/milk products. Further investigation is needed to ascertain whether these reads indicate actual LSDV infections, environmental uptake, or cross-reactivity with related viral sequences.

## 1. Introduction

Lumpy skin disease virus (LSDV) is a double-stranded DNA virus belonging to the Capripoxvirus genus of the *Poxviridae* family. Within this genus are two other virus species: sheeppox and goatpox. LSDV exhibits a strong host specificity and causes disease mainly in *Bos indicus* and *Bos taurus* breeds and water buffalo (*Bubalus bubalis*).

Arthropod vectors primarily transmit LSDV. Studies have demonstrated that *Aedes aegypti* mosquitoes^1^, *Stomoxys calcitrans*^2^, and *Haematopota spp*. biting flies can mechanically transmit LSDV^3^. Additionally, other mosquitoes such as *Culex mirificens* and *Aedes natrionus*^4^, flies like *Biomyia fasciata*, *Culicoides*, and ticks such as *Riphicephalus appendiculatus* and *Amblyomma hebraeum*, are likely to contribute to virus transmission in natural settings^5^. Contact with infected animals is considered to have a minor role in virus transmission^6^. Some studies suggest non-vector transmission such as ingesting contaminated feed and water with infected saliva^7,8^.

LSDV is not a zoonotic virus as there is no evidence to suggest that it can infect humans or cause disease in them. However, in this study, we observed the presence of LSDV reads in the upper respiratory tract (URT) microbiome of humans. The Nasopharyngeal (NP) and Oropharyngeal (OP) samples belong to different locations in the central Indian region of Vidarbha (Maharashtra). These samples were collected as part of the Central government’s SARS-CoV-2 genome surveillance program. These samples were subjected to non-targeted metagenomics to understand the microbial diversity of the samples. The collection period of samples coincides with the period of LSDV circulation in the region. While analyzing the data, the LSDV reads appeared in the samples.

This observation suggests the possibility of non-vector-borne transmission of LSDV. Despite LSDV not infecting humans, extensive bovine-human contact in India through activities such as agriculture, dairy production, and religious rituals raises concerns about the transmission of other diseases from cattle to humans. While LSDV itself isn’t zoonotic, other diseases carried by cattle could pose risks to humans. There could also be a possibility of humans passively transmitting LSDV to cattle.

## 2. Materials and Methods

### 2.1. Sample Collection

NP-OP swab samples were collected in Viral Transport Medium (VTM) by trained paramedical staff at various primary healthcare centres and SARS-CoV-2 molecular testing laboratories from 5 districts of Vidarbha region during March-April 2023. The samples were then tested for SARS-CoV-2 using qRT-PCR. Positive samples ≤ 25 cycle threshold value were selected and transported for SARS-CoV-2 whole genome sequencing (WGS), aliquots of these positive samples were preserved at -80°C. 48 (n) aliquots of these positive samples were selected for whole-genome metagenomics (WGMG) sequencing. The median (interquartile range [IQR]) age was 36 (65 to 16) years, and 24 (50%) of the samples were from females. Location data of every sample was collected and classified as urban and rural households, and the urban-rural distribution of samples stands at 20 (41.6%) and 28 (58.3%) respectively **(Supplementary data)**.

### 2.2. DNA extraction, Library preparation, & Metagenomic Sequencing

DNA extraction was performed with QIAamp DNA Microbiome Kit (51704) per the kit’s protocol. Post-DNA extraction, the Preliminary quality control was done using Qubit to determine the concentration (ng/ul) and Nanodrop (A260/280, A260/230) to determine the purity of extracted metagenomic DNA. Library preparation was done with QIAseq FX DNA Library preparation kit^9^ per the kit’s protocol. The sequencing platform utilised was NextSeq550, employing a 2x150 chemistry high-output 300-cycle kit.

### 2.3. Data Analysis

Fastq files generated after sequencing were analyzed using the Chan Zuckerberg ID (v8.3)^10^, a cloud-based bioinformatic data analysis software. The following steps were involved in the data analysis using Chan Zuckerberg ID (CZid)

#### 2.3.1 Sequencing Data QC

External RNA Controls Consortium (ERCC) sequences were removed using Bowtie2^11^. Sequencing adapters, short reads, sequences with low quality, and low complexity regions were filtered out using a customized version of fastp. Specifically, bases with quality scores below 17, reads shorter than 35 bp, sequences containing low complexity regions exceeding 40%, and sequences with more than 15 undetermined bases (Ns) were excluded.

#### 2.3.2. Host Filtering

Human sequences were removed through Bowtie2^11^ and HISAT2 alignments against reference human genomes. Only one representative read is retained for sequences that are 100% identical based on the first 70 bp, using CZID-dedup. Host reads were filtered out using the Spliced Transcripts Alignment to A Reference (STAR) algorithm. Duplicate, low-quality, and low-complexity sequences were removed in this process. The non-human reads were then aligned to the NCBI nucleotide and protein database (NCBI Index Date: 2021-01-22) using GSNAPL and RAPSearch, respectively.

#### 2.3.3. Alignment

Host-filtered reads were aligned to the NCBI nucleotide (NT) database using Minimap2^12^ and the NCBI protein (NR) database using the Diamond tool^13^. The hits from Minimap2 and Diamond were assigned their corresponding accession numbers. Combined taxon counts were generated by using Gsnap and Rapsearch.

#### 2.3.4. Assembly

To improve sensitivity in mapping, short reads are de novo assembled using SPADES^14^ to generate longer contigs. However, SPADES output lacks information on individual contigs associated with each read. To address this, bowtie2 maps original reads back to assembled contigs, restoring the link between reads and contigs. This process utilizes a Bowtie2 database constructed from contigs. BLAST analysis is then conducted separately for contigs, initially against the NT-BLAST database generated from short-read alignments to NCBI NT using GSNAP, followed by NR-BLAST against NCBI NR using Rapsearch2.

### 2.4. Threshold for Viral Reads Detection

Detection of virus reads is challenging in whole genome metagenome data due to their low abundance, lower genome size, and high genomic diversity. Therefore, selecting the threshold criteria is crucial for reliable detection. Grundy et al. (2023)^15^ experimentally established that 2.4 reads per million (rPM) is the optimum threshold for viral pathogen detection in a metagenomics dataset. We, however used the more conservative filters NT rPM >= 10 and NT L (alignment length) >=35. We utilized the NT and NR sequence filters to mitigate the risk of false identifications in the NCBI database due to misalignment or misannotation.

## 3. Results

### 3.1. Sequencing Data Quality Control (QC)

The data set consists of 48 NP-OP samples. On average (mean ± SD), these samples had 7,258,602 (7.26 million) ± 1,540,000 reads, of which 1,777,507 (1.77 million) ± 5,684,09 reads (25.63 % ± 10.17 %) were identified as nonhuman reads (passed filters). (**Figure 1**) The average minimum and maximum insert sizes were 35 and 557, respectively. The Average Duplicate Compression Ratio (DCR) of data was 1.01 (for metagenomics without enrichment, a DCR value less than 2 is ideal)^16^. Overall, 81% of reads passed the QC following filtering with fastp to eliminate low-quality bases, short reads (less than 35 base pairs in length), and reads with low complexity. (**Figure 2**)

**Figure 1:**
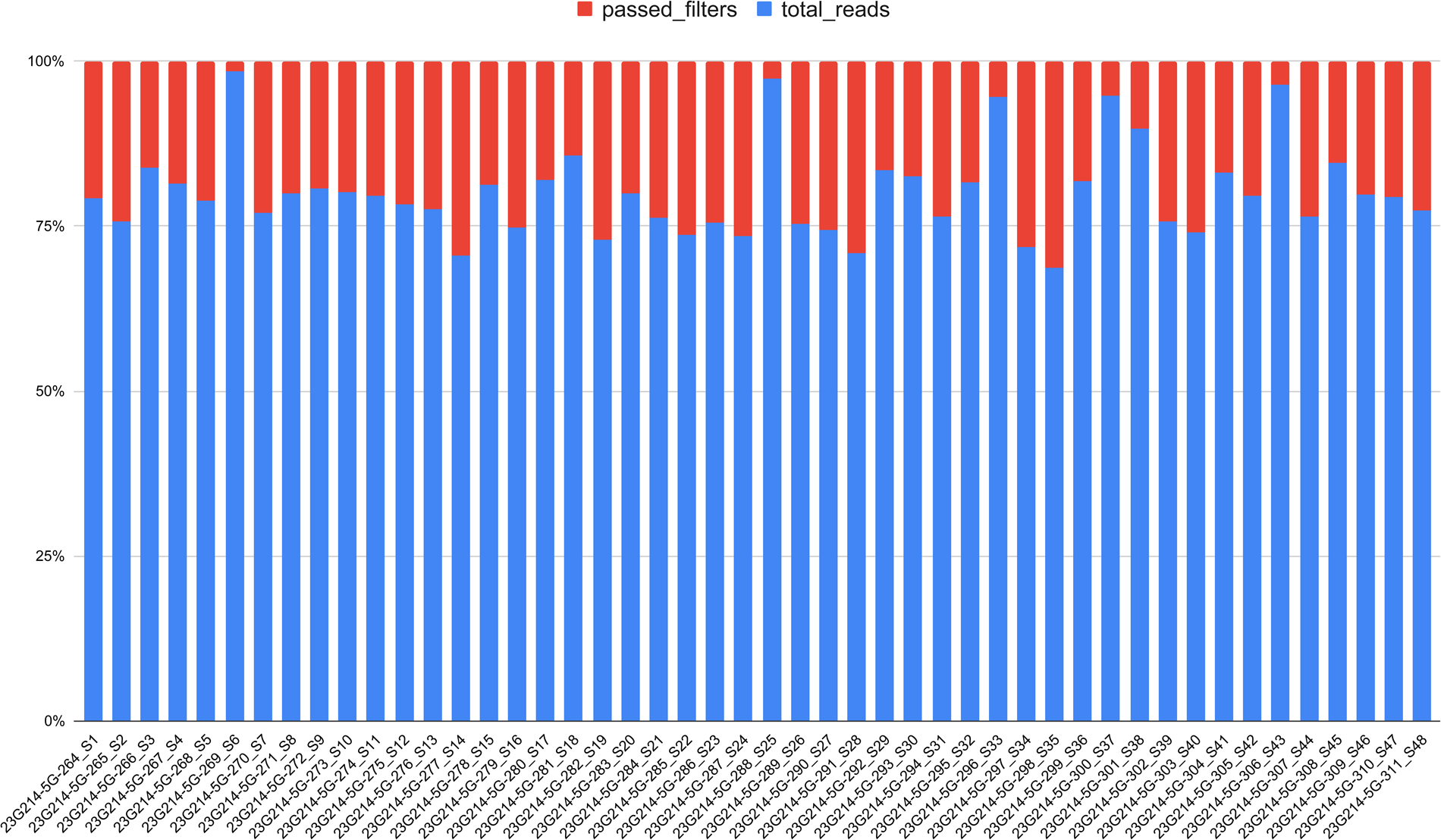
Distribution of total reads and nonhuman reads in the dataset. Each sample, on average, contained approximately 7,258,602 reads, with 25.6% identified as nonhuman reads after passing filters.

**Figure 2:**
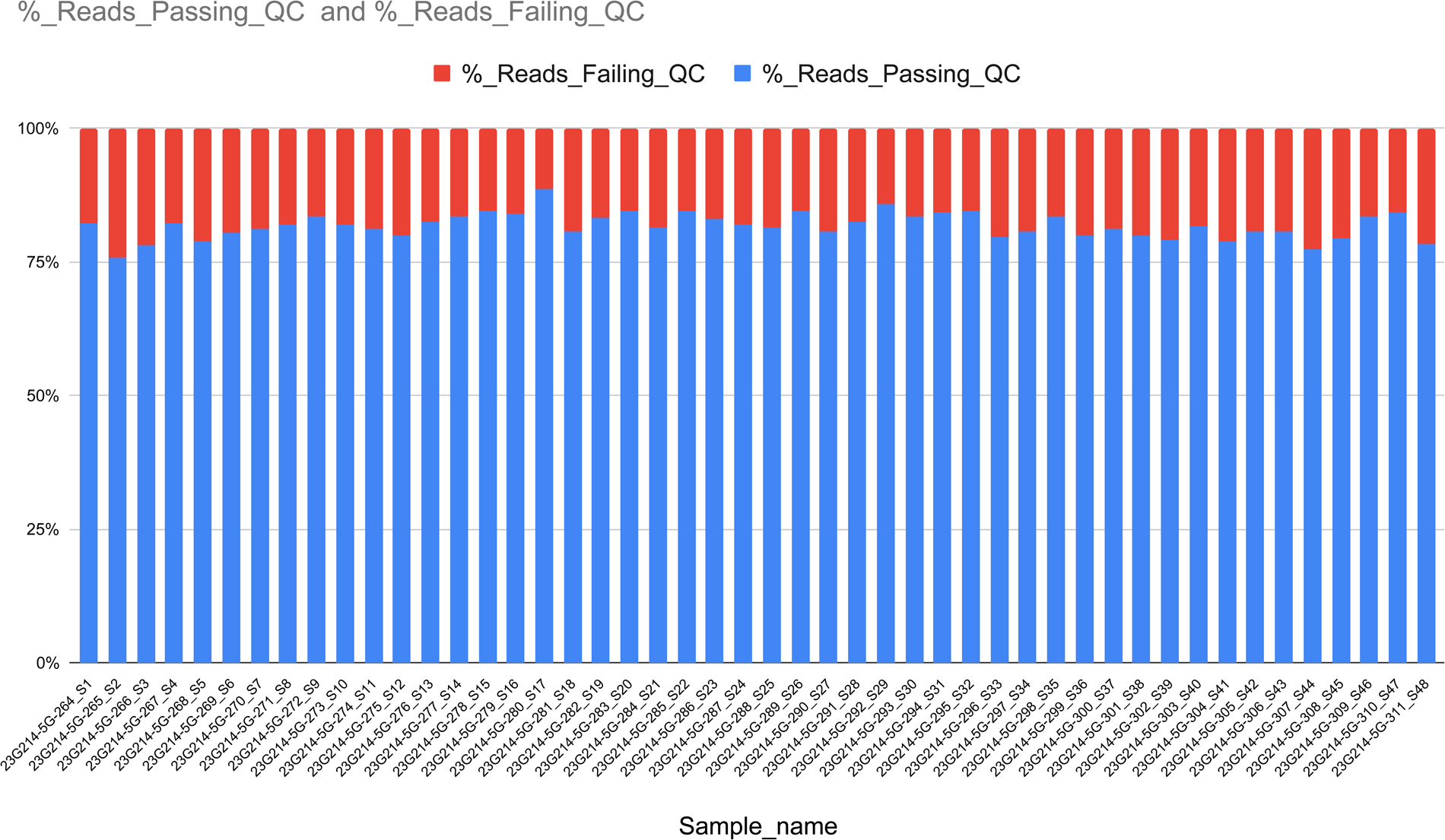
Quality control (QC) results showing the percentage of reads passing filtering with fastp, eliminating low-quality bases, short reads (<35 base pairs), and reads with low complexity. Overall, 81% of reads passed the QC process.

### 3.2. Detection of Viral Reads in the Dataset

After applying specific thresholds for the detection of viral reads in the metagenomic data, including Nucleotide reads per million (NT rPM) >= 10, Nucleotide alignment length in base pairs (NT L) >= 35, and Nucleotide (NT) and Protein (NR) Z Scores >= 1 against a background model of (water only), the analysis revealed the presence of Lumpy Skin Disease Virus (LSDV) reads in 12 out of 48 (25%) samples (**Figure 3**). (sample 23G214-5G-294_S21 was not considered as it had no aligned contigs with the LSDV reference genome KX683219.1) However, it showed 76% coverage and 2.2x depth when aligned to LSDV strain EGPT75 EEV126 gene, partial cds (MH639086.1).

**Figure 3:**
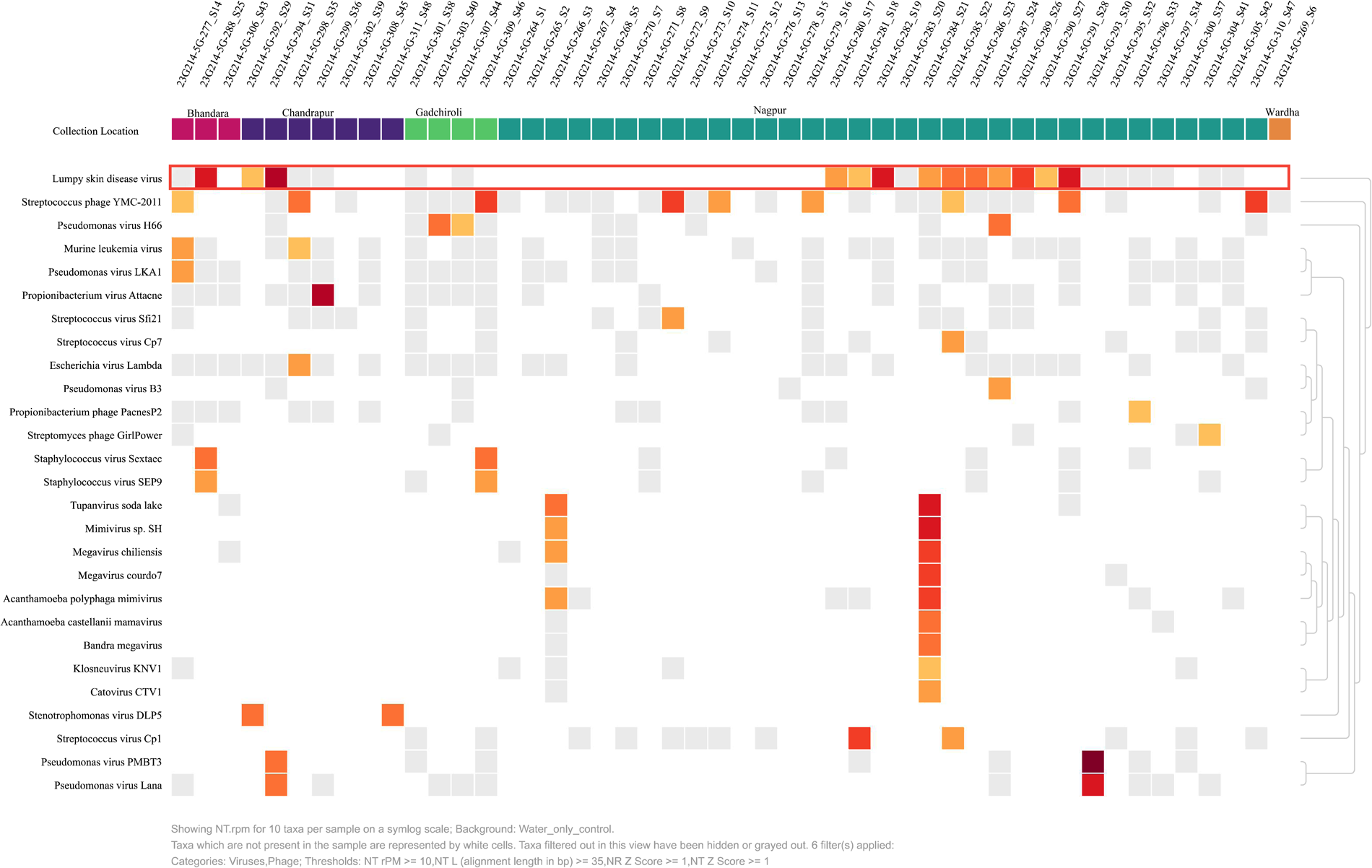
Detection of Lumpy Skin Disease Virus (LSDV) reads in metagenomic samples after applying specific thresholds for viral read detection.

The average alignment length of the reads is 746 ± 231 bases. However, these samples showed a low coverage and sequencing depth for LSDV reads. Samples had 0.6 ± 1.12 average (Coverage Depth X) and 0.87 ± 0.4 (Coverage Breadth %) (**Table 1**)

**Table 1:** Summary of Aligned Contigs, Aligned Loose Reads, Coverage Depth X Coverage Breadth %, and Max Alignment Length of samples containing Lumpy Skin Disease Virus (LSDV) reads.

| Sample | NCBI Reference | Reference Length | Aligned Contigs | Aligned Loose Reads | Coverage Depth X | Coverage Breadth % | Max Alignment Length | Avg. Mismatched % | % Matched |
| --- | --- | --- | --- | --- | --- | --- | --- | --- | --- |
| 23G214-5G-280_S17 | AF409137.1 | 150,793.00 | 1.00 | 0.00 | 0.03 | 0.40 | 617.00 | 0.20 | 99.80 |
| 23G214-5G-281_S18 | AF325528.1 | 150,773.00 | 2.00 | 3.00 | 0.07 | 1.00 | 961.00 | 0.60 | 99.40 |
| 23G214-5G-282_S19 | KY702007.1 | 150,661.00 | 2.00 | 1.00 | 0.23 | 1.00 | 722.00 | 0.40 | 99.60 |
| 23G214-5G-285_S22 | MT130502.2 | 146,159.00 | 2.00 | 1.00 | 0.07 | 1.00 | 722.00 | 0.00 | 100.00 |
| 23G214-5G-286_S23 | MN072619.1 | 150,663.00 | 1.00 | 3.00 | 0.06 | 0.80 | 961.00 | 0.20 | 99.80 |
| 23G214-5G-287_S24 | MH893760.2 | 150,751.00 | 1.00 | 0.00 | 0.01 | 0.20 | 331.00 | 0.30 | 99.70 |
| 23G214-5G-288_S25 | AF409137.1 | 150,793.00 | 4.00 | 13.00 | 3.00 | 1.50 | 961.00 | 0.20 | 99.80 |
| 23G214-5G-289_S26 | MN072619.1 | 150,329.00 | 2.00 | 5.00 | 0.35 | 1.10 | 961.00 | 0.00 | 100.00 |
| 23G214-5G-290_S27 | KX894508.1 | 150,562.00 | 2.00 | 2.00 | 0.02 | 0.60 | 348.00 | 0.00 | 100.00 |
| 23G214-5G-291_S28 | MN995838.1 | 150,721.00 | 2.00 | 2.00 | 0.50 | 0.90 | 772.00 | 0.00 | 100.00 |
| 23G214-5G-292_S29 | KX683219.1 | 150,663.00 | 1.00 | 1.00 | 0.02 | 0.50 | 644.00 | 0.20 | 99.80 |
| 23G214-5G-294_S31 | AF409137.1 | 150,793.00 | 4.00 | 14.00 | 3.00 | 1.50 | 961.00 | 0.20 | 99.80 |
| Mean |  | 150,305.08 | 2.00 | 3.75 | 0.61 | 0.88 | 746.75 | 0.19 | 99.81 |
| SD |  | 1,312.33 | 1.04 | 4.77 | 1.12 | 0.40 | 231.72 | 0.18 | 0.18 |

### 3.3. Consensus Genome

A consensus genome derived from samples containing LSDV (Lumpy Skin Disease Virus) reads would represent a composite sequence assembled from the overlapping sequences obtained from the dataset. The sample with the highest rPM value (1440)was selected to construct the consensus genome. Czid’s consensus genome feature for viral reads was used to create the consensus genome of LSDV ^17^. The consensus genome was built against the LSDV reference sequence (AF409137.1). The consensus genome showed 27.7% GC content, with 2271 informative nucleotides covering 1.5% of the LSDV genome (**Figure 4**).

**Figure 4:**
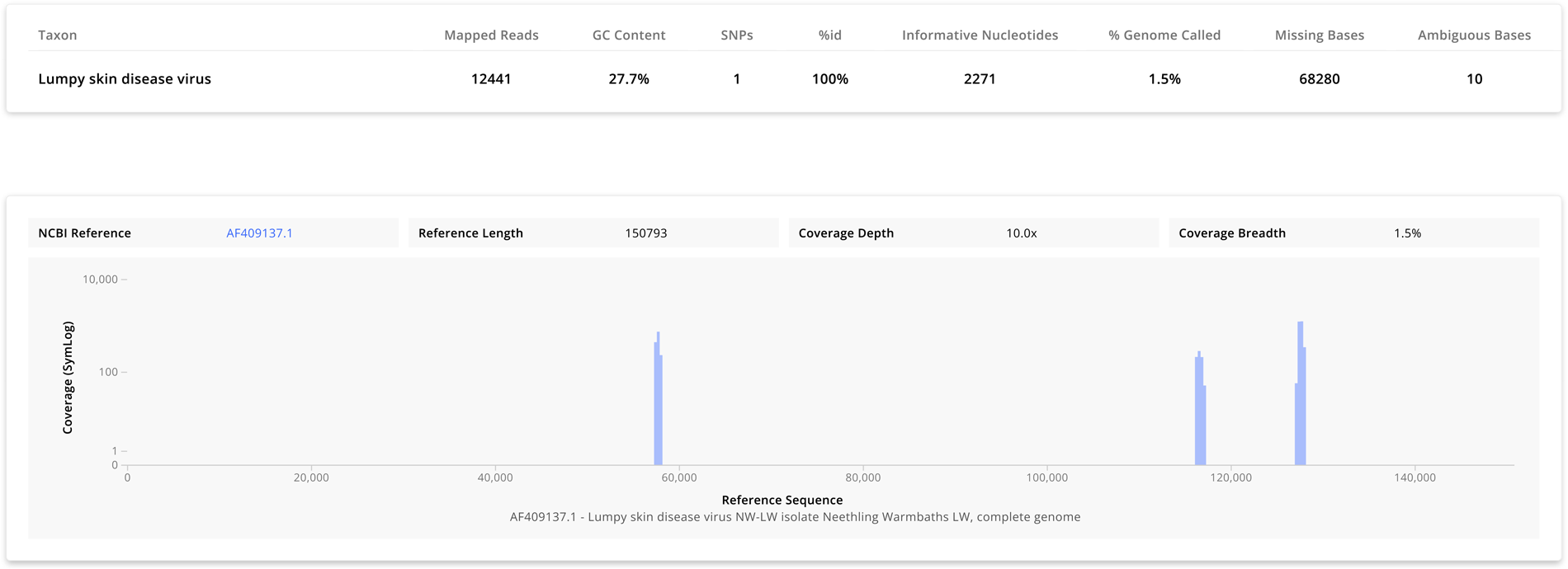
Consensus genome analysis against the LSDV reference sequence (AF409137.1) showing 27.7% GC content and 2271 informative nucleotides, covering 1.5% of the LSDV genome.

### 3.4. General Relationships among the Samples

As the selected samples have contigs covering a small portion of the reference sequence of LSDV (<25%) therefore, the data is not sufficient for constructing a phylogenetic tree^18^; instead, we used Czid’s web-based pipeline to create a pairwise distance matrix for understanding the relationships between identified taxa (LSDV) in the samples. The distance matrix depicts Mash-like distances between samples, calculated using the Jaccard Index (j) based on split kmer matches^19^. Mash-like distances are shown in a pairwise matrix, where each sample is compared to itself and others. Colors represent distance ranges, with dark red squares along the diagonal indicating identical sequences (**Figure 5**).

**Figure 5:**
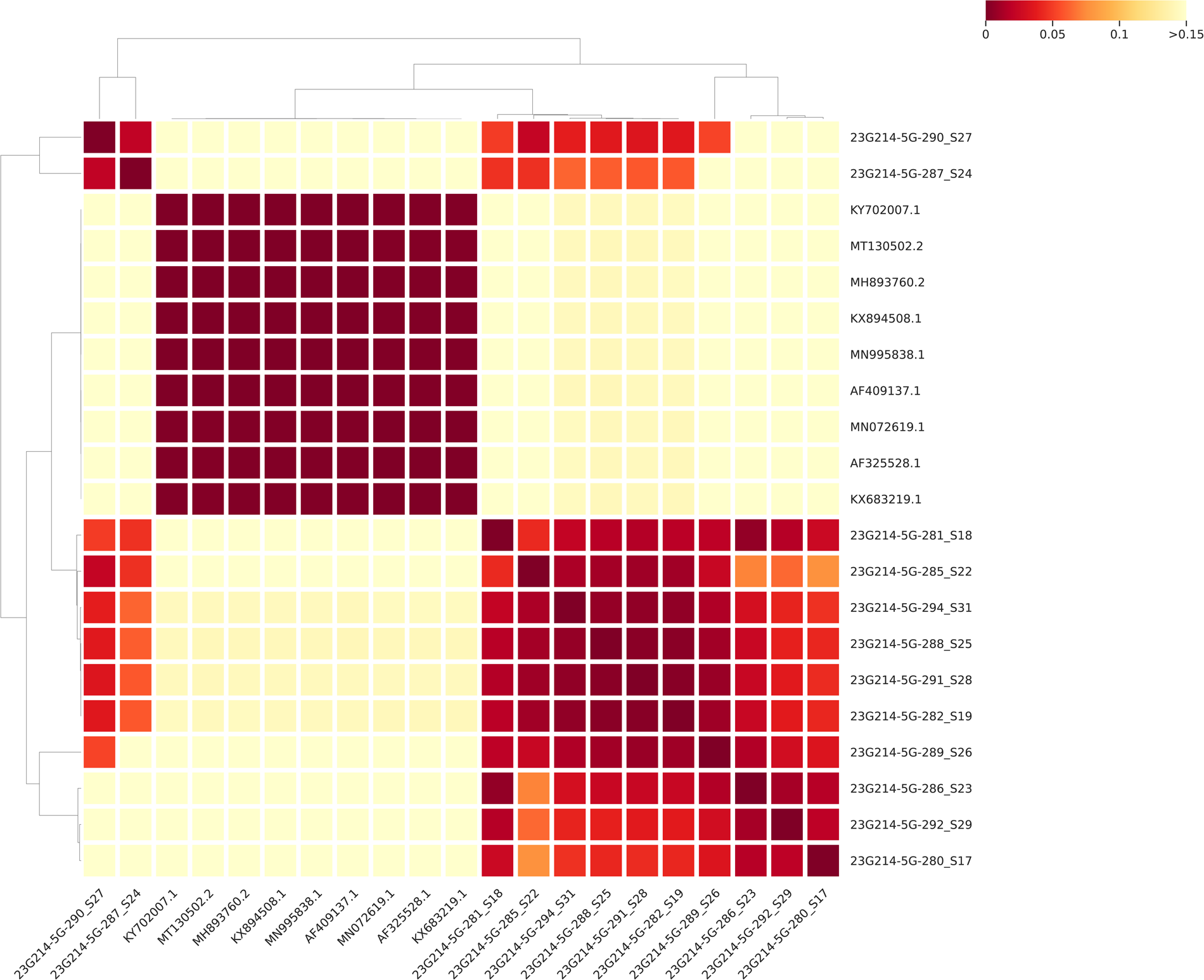
Pairwise Mash-like distance matrix illustrating Jaccard Index (j) based on split kmer matches between samples. Dark red squares along the diagonal indicate identical sequences, with colors representing distance ranges.

### 3.5. Genomic Position and Classification of (LSDV) Contigs

Czid’s coverage visualisation tool was used to determine the location of contigs over the reference genome. The corresponding accession numbers of reference genomes were also annotated. These samples were classified as wild-type (WT) and vaccine strain (V) based on the classifications as reported by Flannery et al. (2021)^20^. The table also includes information on each sample’s total number of contigs and specifies its genomic position over the reference genome. It was observed that the contigs aligned to some specific regions of the reference genomes; these regions are 55109 to 57997 bp, 115916 to 117098 bp, 116144 to 117104 bp, and 124934 to 127961 bp. (**Table 2**) This observation is indicative of a probable common source of the LSDV reads.

**Table 2:** Details of LSDV Samples: Classification of Vaccine Strain (V) / Wild type Strain (WT), Contig Details, and their Genomic Position.

| S. No. | Sample | NCBI<br>Accession No. | Name of Accession | Vaccine<br>Strain (V) /<br>Wild type<br>Strain (WT) | Total<br>Contigs | Contig 1<br>Position (bp) | Contig 2<br>Position<br>(bp) | Contig 3<br>Position<br>(bp) | Contig 4<br>Position<br>(bp) |
| --- | --- | --- | --- | --- | --- | --- | --- | --- | --- |
| 1 | 23G214-5G-280_S17 | AF409137.1 | Lumpy skin disease virus NW-LW isolate<br>Neethling Warmbaths LW, complete genome | WT | 1 |  | 116306-116922 |  |  |
| 2 | 23G214-5G-281_S18 | AF325528.1 | Lumpy skin disease virus NI-2490 isolate<br>Neethling 2490, complete genome | WT | 2 | 57401-57699 | 116138-117098 |  |  |
| 3 | 23G214-5G-282_S19 | KY702007.1 | Lumpy skin disease virus isolate<br>SERBIA/Bujanovac/2016, complete genome | WT | 2 |  | 115916-116876 |  | 127172-127893 |
| 4 | 23G214-5G-285_S22 | MT130502.2 | Lumpy skin disease virus strain Neethling-RIBSP vaccine, partial genome | V | 2 | 55109-55703 |  |  | 124934-125655 |
| 5 | 23G214-5G-286_S23 | MN072619.1 | Lumpy skin disease virus isolate Kenya,<br>complete genome | WT | 1 |  | 115916-116876 |  |  |
| 6 | 23G214-5G-287_S24 | MH893760.2 | Lumpy skin disease virus strain<br>LSDV/Russia/Dagestan/2015, complete<br>genome | WT | 1 | 57395-57725 |  |  |  |
| 7 | 23G214-5G-288_S25 | AF409137.1 | Lumpy skin disease virus NW-LW isolate<br>Neethling Warmbaths LW, complete genome | WT | 4 | 57403-57997 | 116144-116475 | 116634-117104 | 127240-127961 |
| 8 | 23G214-5G-289_S26 | MN072619.1 | Lumpy skin disease virus isolate Kenya,<br>complete genome | WT | 2 |  | 115916-116876 |  | 127007-127728 |
| 9 | 23G214-5G-290_S27 | KX894508.1 | Lumpy skin disease virus isolate<br>155920/2012, complete genome | WT | 2 | 57404-57739 |  |  | 127252-127599 |
| 10 | 23G214-5G-291_S28 | MN995838.1 | Lumpy skin disease virus isolate pendik,<br>complete genome | WT | 2 | 57361-57955 |  |  | 127196-127917 |
| 11 | 23G214-5G-292_S29 | KX683219.1 | Lumpy skin disease virus strain KSGP 0240,<br>complete genome | V | 1 |  | 116256-116896 |  |  |
| 12 | 23G214-5G-294_S31 | AF409137.1 | Lumpy skin disease virus NW-LW isolate<br>Neethling Warmbaths LW, complete genome | WT | 4 | 57403-57997 | 116199-116244 | 116144-117104 | 127240-127961 |

### 3.6. CZid pipeline compared with Centrifuge

CZid uses the Genomic Short-read Nucleotide Alignment Program GSNAP^21^ to align the reads with the reference genome using the NCBI index. Centrifuge^22^ employs an indexing strategy derived from the Burrows-Wheeler transform (BWT) and the Ferragina-Manzini (FM) index for efficient metagenomic classification. We compared the results from CZid (rPM) and Centrifuge (Unique Reads) to avoid any pipeline-dependent biases; both results followed a similar trend, as shown in (**Figure 6**) detailed results are shown in (**Table 3**)

**Figure 6:**
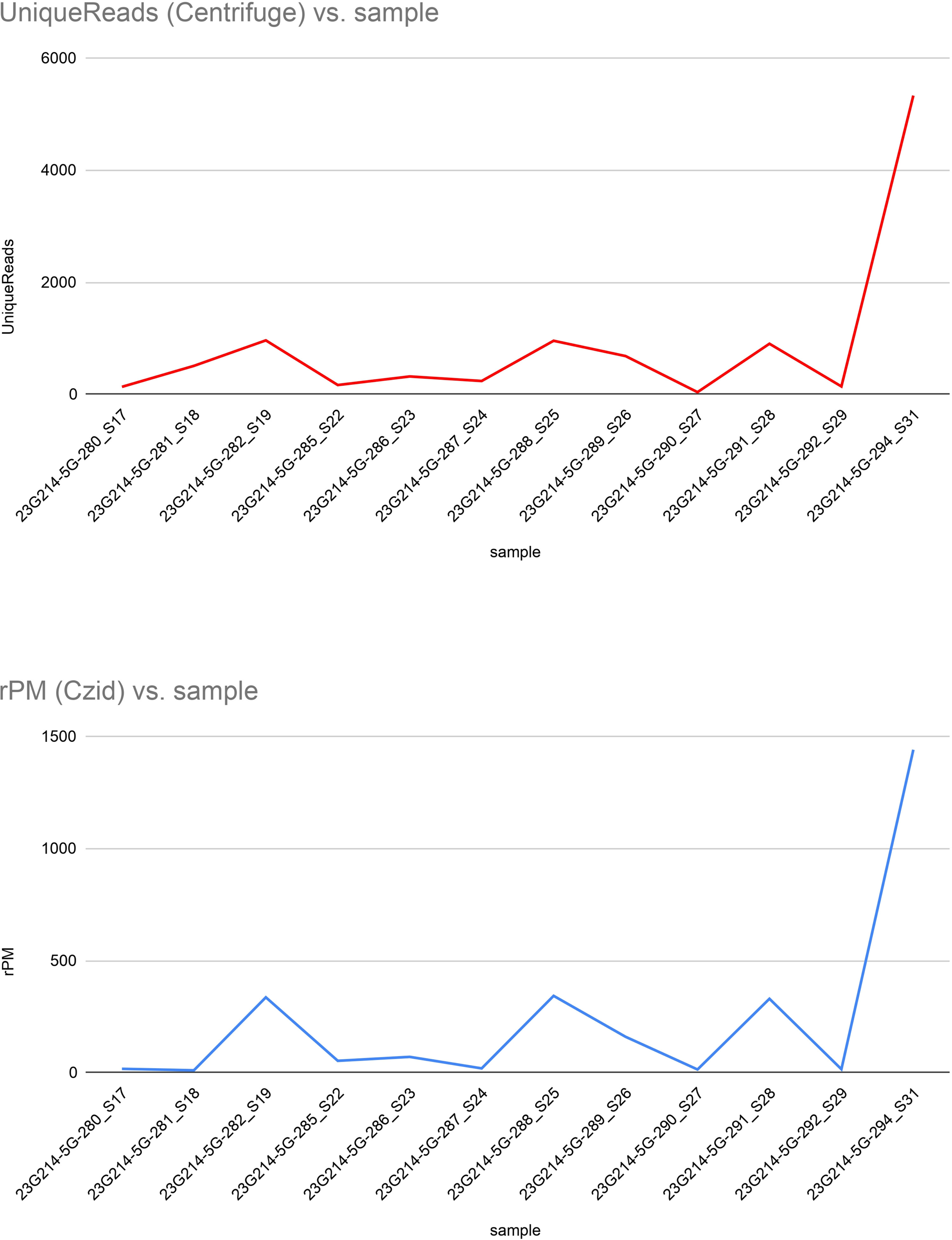
Comparison of CZid (rPM) and Centrifuge (Unique Reads) pipeline results, demonstrating a consistent trend across both methods.

**Table 3:** Summary of Taxon names, taxon IDs, taxonomic ranks, number of reads, number of unique reads, abundance, and relative abundance (reads per million) of samples with LSDV reads as determined by Centrifuge and CZid.

| S. No. | sample | Taxon name (Centrifuge) | taxID (Centrifuge) | taxRank (Centrifuge) | numReads (Centrifuge) | numUniqueReads (Centrifuge) | Abundance (Centrifuge) | rPM (CZid) |
| --- | --- | --- | --- | --- | --- | --- | --- | --- |
| 1 | 23G214-5G-280_S17 | Lumpy skin disease virus NI-2490 | 376849 | leaf | 154 | 134 | 0.000272504 | 17.1864 |
| 2 | 23G214-5G-281_S18 | Lumpy skin disease virus NI-2490 | 376849 | leaf | 538 | 508 | 0.049054 | 10.5188 |
| 3 | 23G214-5G-282_S19 | Lumpy skin disease virus NI-2490 | 376849 | leaf | 1096 | 963 | 0.0503631 | 336.258 |
| 4 | 23G214-5G-285_S22 | Lumpy skin disease virus NI-2490 | 376849 | leaf | 185 | 166 | 0.00782118 | 52.9468 |
| 5 | 23G214-5G-286_S23 | Lumpy skin disease virus NI-2490 | 376849 | leaf | 376 | 319 | 0.0106748 | 70.5397 |
| 6 | 23G214-5G-287_S24 | Lumpy skin disease virus NI-2490 | 376849 | leaf | 279 | 238 | 0.00439487 | 19.1551 |
| 7 | 23G214-5G-288_S25 | Lumpy skin disease virus NI-2490 | 376849 | leaf | 1083 | 956 | 0.195623 | 342.871 |
| 8 | 23G214-5G-289_S26 | Lumpy skin disease virus NI-2490 | 376849 | leaf | 751 | 683 | 0.0185272 | 160.216 |
| 9 | 23G214-5G-290_S27 | Lumpy skin disease virus NI-2490 | 376849 | leaf | 49 | 39 | 0.0120165 | 14.2801 |
| 10 | 23G214-5G-291_S28 | Lumpy skin disease virus NI-2490 | 376849 | leaf | 1024 | 903 | 0.0321114 | 329.775 |
| 11 | 23G214-5G-292_S29 | Lumpy skin disease virus NI-2490 | 376849 | leaf | 159 | 141 | 0.000831785 | 15.5906 |
| 12 | 23G214-5G-294_S31 | Lumpy skin disease virus NI-2490 | 376849 | leaf | 5956 | 5332 | 0.0544535 | 1440.89 |

## 4. Discussion

In this study, we have reported the detection of LSDV reads in the human upper respiratory tract microbiome. 48 NP-OP swab samples from 5 districts of Maharashtra were randomly selected for this study; these samples were collected from different healthcare centers and testing labs for SARS-CoV-2 genome surveillance. These samples were then subjected to enrichment-free whole-genome metagenomics (WGMG) sequencing to investigate upper respiratory tract microbiome composition.

The generated data set consists of 7.25 million reads on average, with an average of 25% of reads passing the host filters. The detection of LSDV reads was consistently observed among the samples; 20 out of 48 samples were detected with LSDV reads before applying the selection threshold criteria of NT rPM >= 10 and NT L (alignment length) >=35. After using this criterion, the number of samples with LSDV reads detection decreases to 12 (25%) of the total samples.

It was observed that the LSDV viral contigs showed less coverage with the reference genome. The low coverage of LSDV viral contigs with the reference genome could be attributed to several factors, including generally low biomass NP-OP sampling.^23^ The samples involved in this study were OP-NP swab samples collected in VTM by primary healthcare workers from urban and rural regions of Vidarbha (Central India). Maintaining Cold chain transport is challenging for samples from rural areas; a break in the cold chain may contribute to sample degradation, compromising the coverage during sequencing.

Also, factors like the physical conditions of NP-OP, depth of sampling, the presence of mucous, and the age of the participants may determine the availability of biomass/tissue material while sampling. Metagenomics data generation, especially with low biomass NP-OP swab samples, presents challenges due to limited microbial material, complex microbial communities, and host DNA contamination.^24,25,26^

Another reason for the observed low coverage of LSDV reads could be genetic variability within the viral population that can lead to mismatches between the reference genome and sequenced reads, reducing alignment efficiency and coverage of viral contigs.^27,28^

As the contigs covered only a limited portion of the reference sequence of LSDV in the selected samples, this prevented the construction of a phylogenetic tree. Therefore, we used Czid’s web-based pipeline, which allowed us to create a pairwise distance matrix. This pipeline used the split kmer analysis (SKA) method to determine the degree of relatedness among the samples.SKA enables quick comparison and alignment of shorter genome fragments, which makes it well-suited for surveillance and outbreak analyses. It can create split kmer files from fasta or fastq formats, cluster them, align them with a reference sequence, and offer various comparative and summary metrics. SKA has demonstrated accuracy with Illumina sequencing data.^29^ Furthermore, a comparison between CZid and Centrifuge pipelines demonstrated similar trends in results, indicating consistency across different analysis methods.

The detection of LSDV reads in the human upper respiratory tract microbiome poses intriguing questions and warrants further investigation. While LSDV is traditionally associated with bovines and causes significant economic losses in the agricultural sector, its presence in the human respiratory tract microbiome indicates a potential uptake by humans from the environment. LSDV is primarily transmitted through arthropod vectors and direct contact with infected animals, thus making its presence in the human upper respiratory tract unexpected. Out of 12 samples with LSDV reads, seven belong to rural areas, and five belong to Urban areas. LSDV reads in the URT microbiome could also be attributed to frequent contact between humans and livestock in India.^30^

Another plausible explanation for LSDV reads detection in URT could be traced to consuming milk from infected bovines. Bedkovic (2018) ^31^ reported the detection of LSDV viral particles in Milk. These viral particles in milk could either be from naturally infected or animals vaccinated with Live, Attenuated Vaccines, as in the case of LSDV. DNA from LSDV wildtype and vaccine strains were also detected in the milk and body fluids of vaccinated animals.^31,32^ Our data also showed the detection of contigs from both wild-type and Vaccine strains of LSDV. These contigs were observed to be consistently aligning to specific locations on the reference genome, this observation is suggestive of a common source for LSDV reads. This common source is more likely to be a water source shared by both humans and animals or milk/milk-products from LSDV infected or Vaccinated bovines.

Therefore, it is crucial to investigate whether these reads represent actual LSDV infections in humans, take-up from the environment, or cross-reactivity with related viral sequences. Accurate taxonomic classification and identification of viral sequences require comprehensive reference databases and stringent alignment criteria to minimize false-positive results and ensure the reliability of findings.^33^ Therefore, investigations in this direction further validate the presence of LSDV reads using alternative experimental approaches and studies in controlled environments.

## Acknowledgement

The authors are thankful to CSIR-NEERI for providing funds under project OLP-57 (March 2023 - April 2024) for conducting this study

## Data availability statement

The data for the samples in this study is available as **“Viral_Metagenome_Covid”** on CZid.

## Author declaration

The author assures that the research has followed all ethical guidelines and received approvals from the Institutional Ethics Committee for Research on Human Subjects (IEC) of CSIR-NEERI, Nagpur-20, India. Necessary consent from patients/participants has been obtained and relevant institutional documentation is archived.

## Confidentiality declaration

Sample IDs **(23G214-5G-264_S1 to 23G214-5G-311_S48)** are masked IDs and cannot be traced to participant details. Precise age of the participants is masked and non-ovelapping age ranges were used.

